# Fusion of Fully Integrated Analog Machine Learning Classifier with Electronic Medical Records for Real-time Prediction of Sepsis Onset

**DOI:** 10.1101/2021.06.24.21259466

**Authors:** Sudarsan Sadasivuni, Monjoy Saha, Neal Bhatia, Imon Banerjee, Arindam Sanyal

**Affiliations:** University at Buffalo, Electrical Engineering, Buffalo, 14260, USA; Emory University, Department of Biomedical Informatics, Atlanta, 30322, USA; Emory University, Department of Radiology, Atlanta, 30322, USA; Emory University School of Medicine, Division of Cardiology, Atlanta, 30322, USA

**Author notes:** these authors contributed equally to this work (Sudarsan Sadasivuni, Monjoy Saha). co-senior authors (Imon Banerjee, Arindam Sanyal).

## Abstract

The objective of this work is to develop a fusion artificial intelligence (AI) model that combines patient electronic medical record (EMR) and physiological sensor data to accurately predict early risk of sepsis. The fusion AI model has two components - an on-chip AI model that continuously analyzes patient electrocardiogram (ECG) data and a cloud AI model that combines EMR and prediction scores from on-chip AI model to predict fusion sepsis onset score. The on-chip AI model is designed using analog circuits for sepsis prediction with high energy efficiency for integration with resource constrained wearable device. Combination of EMR and sensor physiological data improves prediction performance compared to EMR or physiological data alone, and the late fusion model has an accuracy of 93% in predicting sepsis 4 hours before onset. The key differentiation of this work over existing sepsis prediction literature is the use of single modality patient vital (ECG) and simple demographic information, instead of comprehensive laboratory test results and multiple vital signs. Such simple configuration and high accuracy makes our solution favorable for real-time, at-home use for self-monitoring.

## 1 Background

The USA has one of the highest healthcare costs in the world even though it lags behind peer nations in health outcomes. Advances in artificial intelligence (AI) and continuous health surveillance technologies can alleviate some of the healthcare burden through early detection of health issues. Our objective is to develop on-chip AI circuit that will continuously monitor physiological signals and build a fusion model to predict early risk of clinically adverse event to increase the management time and prevent adverse outcome (see Figure 1). In this study, we used sepsis as our use-case since sepsis has high mortality rates around 40% for severe cases after onset^1^, and almost 80% of sepsis patients have onset outside hospital. Real-time, continuous at-home health surveillance leveraging AI can significantly improve outcomes for sepsis by providing early alert to patients and allowing them to seek timely medical intervention which may significantly reduce the mortality rate.

**Figure 1.**
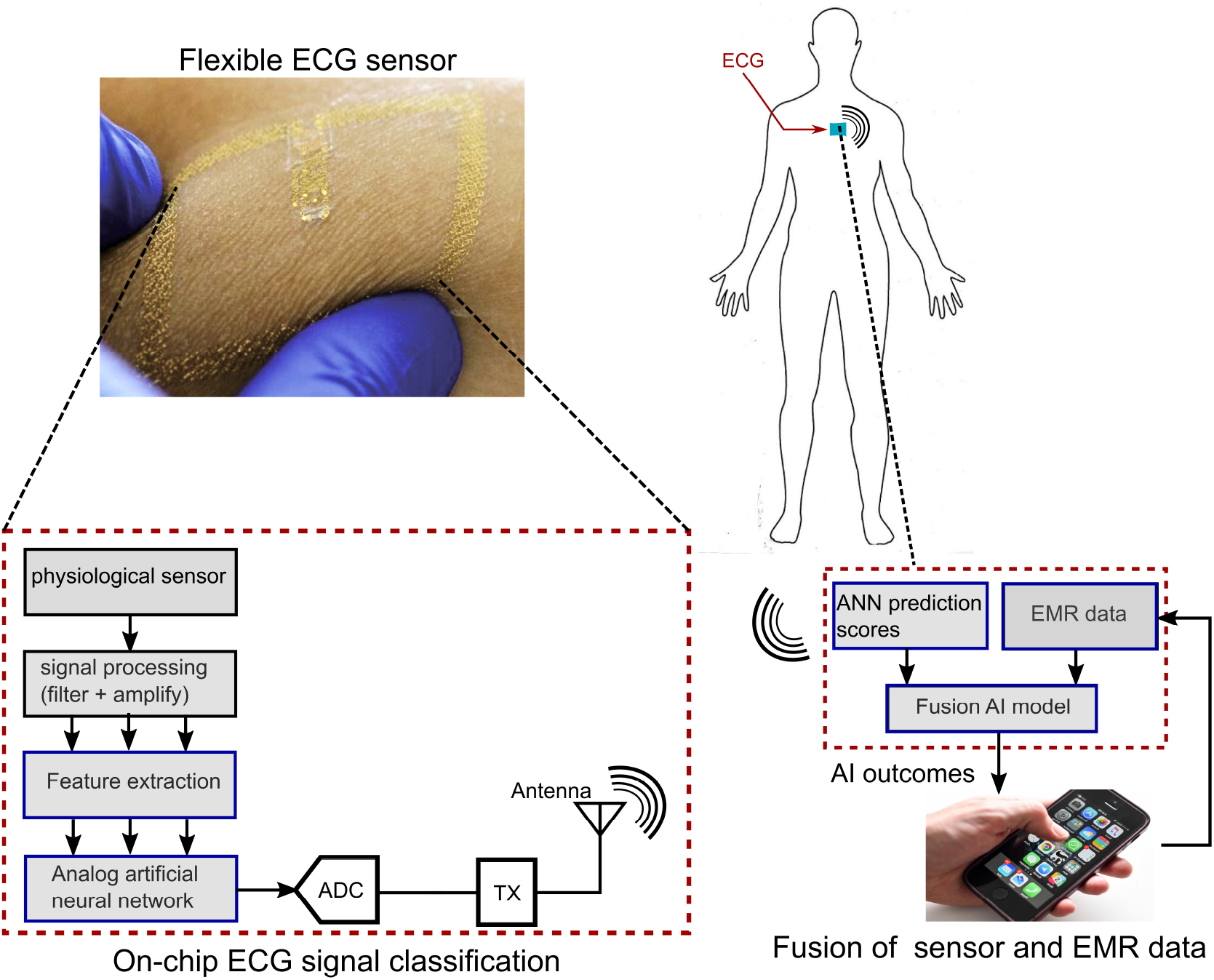
Overview of the proposed paradigm with items outlined in blue representing the tasks in this work

While recent clinical trials, such as the Apple Heart Study^2^, have demonstrated the utility of wearable device for continuous health monitoring, there are several challenges and limitations associated with the current technologies - 1. existing wearable devices lack the ability to integrate electronic medical record (EMR) with sensor data in real-time, even though EMR is known to have a strong influence on health abnormality detection^3–6^; 2. most wearable devices do not have automated inference capability and depend on telemetry and medical experts for actionable inference; 3. the current framework of performing AI analysis in the cloud increases risks of breach in patient data during transmission over the network^7,8^. The proposed work addresses these challenges through a two-step fusion AI framework - an AI circuit that can be integrated with wearable device for performing in-situ analysis of continuous sensor data, and a cloud AI model that performs fusion of demographic data and scores from embedded AI circuit for real-time risk prediction of sepsis onset. Raw patient data collected through wearable device is not transmitted to the cloud; rather only prediction scores of the embedded AI circuit is sent to the cloud for fusion which improves robustness against eavesdropping attacks on the patient’s sensor data.

The proposed two-step *fusion AI solution* partitions the data analysis into ECG and EMR data models (Figure 1). The ECG AI model will be integrated with physiological sensor attached to the patient body, and will continuously analyze sensor data and classify it into sepsis and non-sepsis risk for varying onset time. The prediction score from the on-chip ECG model will be transmitted to the external EMR AI model that will be implemented on cloud processor and use deep-learning techniques to perform fusion of physiological and EMR data for precision health decisions. The user will enter EMR data through a smart-phone application during initial registration, and the AI model will alert the user to abnormal health conditions. Our hypothesis is that integration of sensor and EMR data will significantly improve clinical decision accuracy, while selective transmission will extend un-interrupted patient monitoring span and improve security against unwanted access.

## 2 Results

Experiments in the study were carried out following the World Medical Association (WMA) developed ethical protocol for conducting ethical medical research. The model was trained and tested on 965 unique patients (514 sepsis and 451 non-sepsis) from Emory Healthcare system and the sepsis onset time is defined by Sepsis-3 definition^9^. As a baseline, the prediction results using only single modality models (ECG and EMR separately) have been shown in Table 1.In this experiment with EMR models, we used only patient demographic data (age, gender, race, ethnicity) and co-morbidity information represented in textual format. Out of four different classifier models, random forest classifier with boosting strategy showed the highest classification accuracy 76% with area under the curve (AUC) of 0.86 for identifying sepsis risk with demographics and co-morbidity. The average classification accuracy by rest of the classifiers was around 51% which shows the fact that EMR data alone, even including previous clinical history, is not a good predictor for sepsis prediction task. For our further experiments, we used random forest classifier since it provides the optimal performance on the baseline classification task. The stand-alone ECG model achieved a 95% accuracy for predicting sepsis within 1 hour. While the ECG model achieved high accuracy for short-term prediction, the baseline performance of the ECG model was 86% and 77.5% accuracy for predicting sepsis 4 hours and 6 hours before the onset time which is not optimal for taking early preventive measures for sepsis at-home.

**Table 1.**
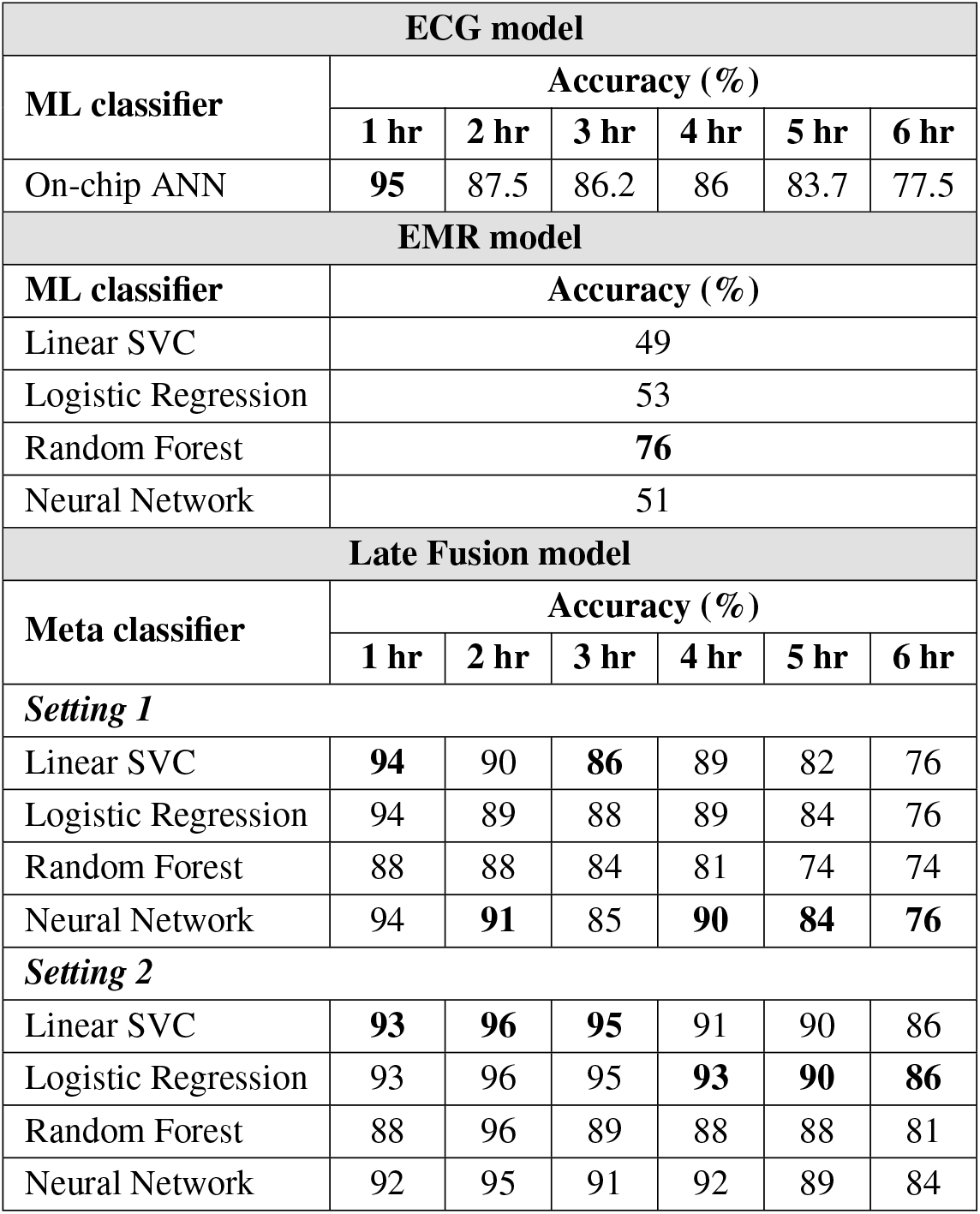
Baseline prediction results using ECG on-chip model, and demographics and co-morbidity data. Late fusion model performance - Setting 1: using demographic and ECG data; Setting 2: using demographic, co-morbidity and ECG data. Optimal performance for every prediction task is highlighted in **bold**.

In order to improve the accuracy for early prediction, we developed a late fusion model where we trained a meta-learner on the prediction scores of on-chip ECG model and EMR models to integrate the information from two data sources. Table 1 show late fusion results. In order to interpret the importance of different data sources, we performed the fusion in two different settings - *setting 1*: late fusion was performed using demographic, and ECG data; *setting 2*: late fusion was performed using demographic, co-morbidity, and ECG data. Late fusion was performed for 1 hour to 6 hours before the sepsis onset time. Figure 2 shows the area under the receiver operating characteristic curves (AUROC) for late fusion model using demographic, co-morbidity, and ECG data at different time steps.

**Figure 2.**
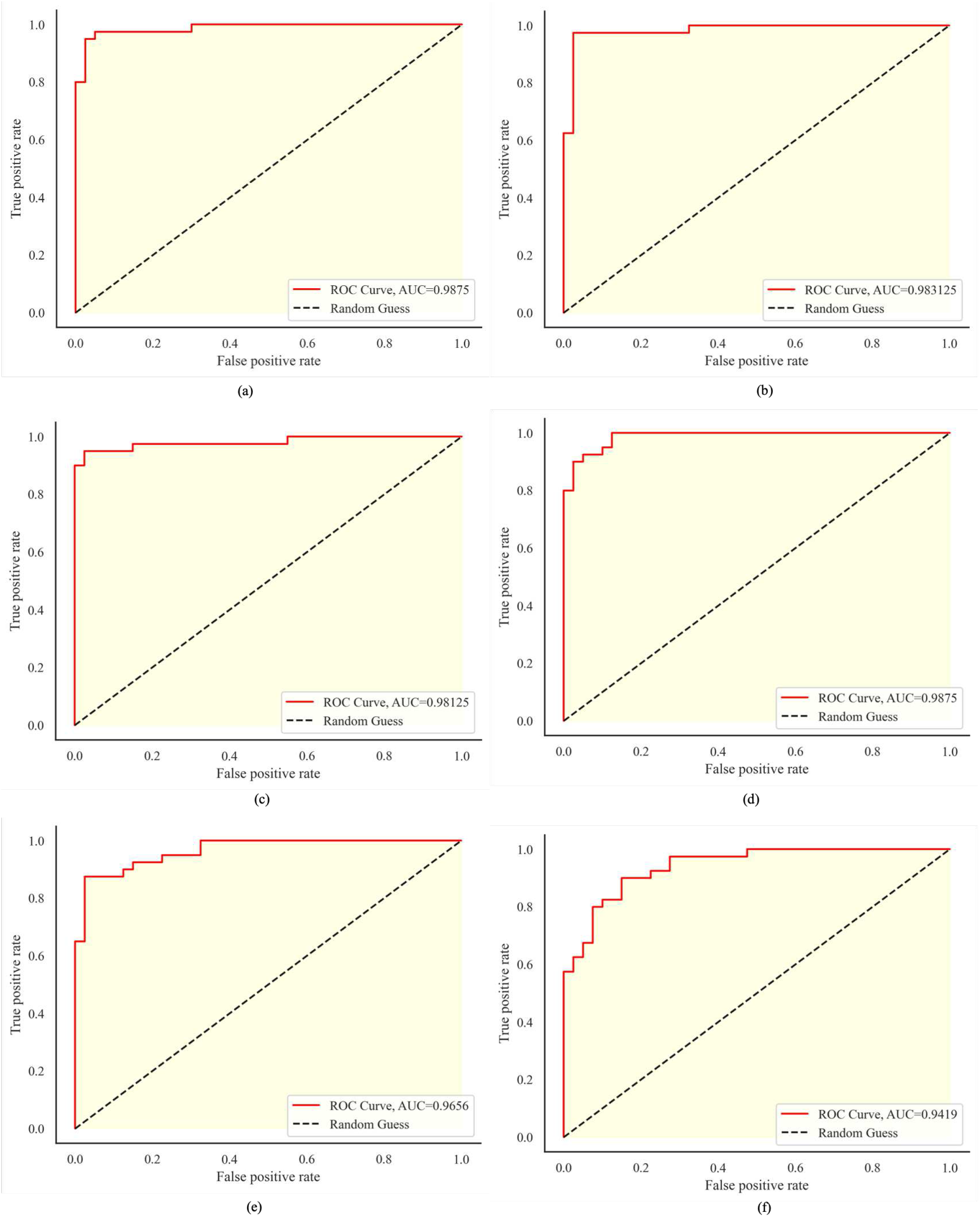
Receiver operating characteristic curve for late fusion using demographic, co-morbidity, and ECG data at different time steps; (a) 1 hr; (b) 2 hrs; (c) 3 hrs; (d) 4 hrs; (e) 5 hrs; (f) 6 hrs

Fusion models in general improved the baseline EMR model performance from 76% to 93% for 1 hour and 86% for 6 hours prediction before the sepsis onset. The baseline EMR model’s AUROC values also improved from 0.86 to 0.99 for 4hrs prediction task (Figure 2). An interesting observation can be drawn from the fusion models’ performance that the prediction of sepsis close to the onset time (e.g. 1 hour) achieved better performance (94%) without the clinical history of the patients while the clinical history combined with demographics and sensor data provides a significant boost in performance for early sepsis prediction - 5 hours - improved from 84% to 90% and 6 hours - improved from 77.5% to 86%. We present details of the performance analysis in the supplemental material.

We designed a graphical mobile application for easy adaptability from the user view-point (Figure 3). After initial registration by the user, the application collects the sensor prediction from the AI model integrated within the sensor patch and collects the demographic and clinical history data from the user. Our prediction fusion algorithm is the core backbone of the application that will compute the risk of sepsis and visualize the risk prediction for 6 different onset times in a dial format.

**Figure 3.**
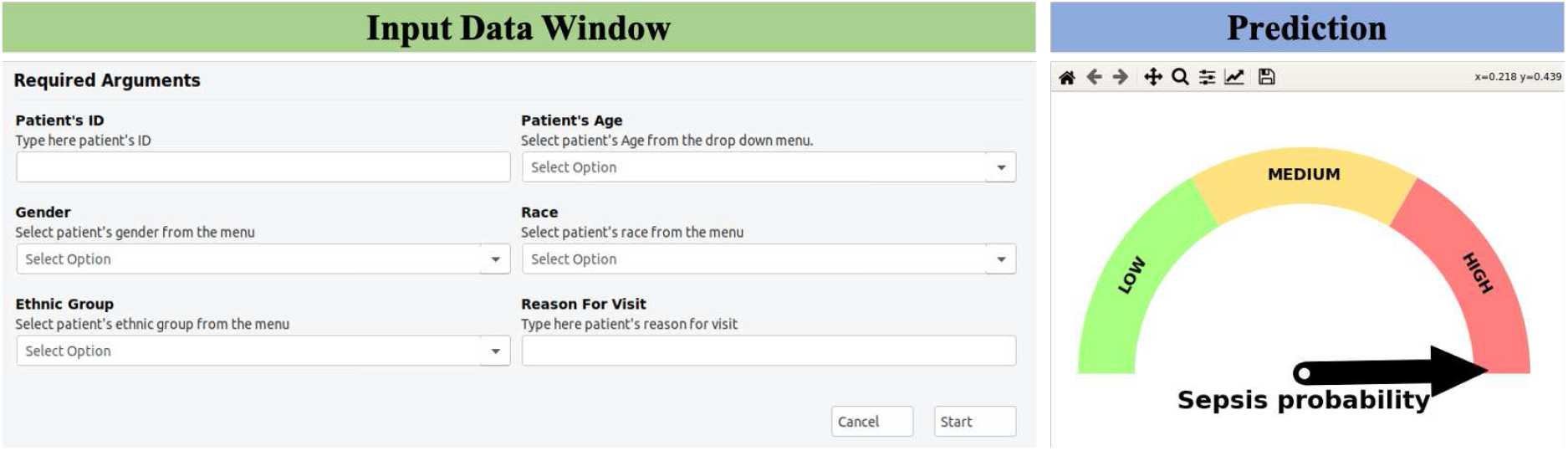
A graphical user interface for sepsis prediction with fusion AI model

## 3 Discussion

In this study, we developed an innovative fusion AI solution for sepsis prediction and demonstrate the performance on varying prediction time-points. Core novelty of this work is - (1) the first on-chip ECG model with fusion AI achieved state-of-the-art accuracy for sepsis prediction; (2) analog in-memory computing (IMC) for high energy efficiency AI model in the ECG wearable. In order to support our novelty claim, we compared our solution with the existing literature in both AI and analog domain in the following subsections.

### 3.1 Comparison with prior works - first on-chip fusion AI model with state-of-the-art accuracy

Table 2 compares this work with state-of-the-art sepsis prediction literature in terms of performance, model architecture, and required data sources. In contrast to prior works that combines patient vitals from multiple modalities, laboratory test results and demographics information, our work uses only patient ECG signal and demographics which makes the proposed solution feasible for real-time, at-home patient monitoring and is a key differentiation over existing works. Our work has high accuracy, sensitivity, specificity and AUROC metrics, and the best prediction performance for time-to-sepsis onset of 4 hours or less. The work by^10^ has higher accuracy, but requires 26 different laboratory test results and 8 different patient vital signs. The work by Wickramaratne et. al.^11^ uses a fusion technique to combine EMR and patient vitals, but requires 11 laboratory test results, 18 culture tests, 6 vital signs and 30 co-morbidities which is only feasible in a hospital setting. None of the existing AI models are implemented within the ECG sensor attached to the patient body for real-time risk inference while preserving the data privacy.

**Table 2.** Comparison with state-of-the-art AI models for sepsis prediction

|  | Model | Performance Metrics |  |  |  |  | Data Source |  |  |
| --- | --- | --- | --- | --- | --- | --- | --- | --- | --- |
|  |  | t<sub>onset</sub> (hrs) | Accuracy | Sensitivity | Specificity | AUROC | Vitals | Lab<sup>1</sup> | Dem.<sup>2</sup> |
| <sup>12</sup>CinC 2019 | LSTM | 4 | 84.5% | — | 0.66 | 0.8 | 8 | 26 | 6 |
| <sup>13</sup>CIBM 2016 | Ensemble | 3 | 82.7% | 0.81 | 0.9 | 0.83 | 9 | 0 | 0 |
| <sup>14</sup>CinC 2019 | Random forest | 6 | 74.6% | — | — | 0.63 | 8 | 26 | 6 |
| <sup>15</sup>CinC 2019 | Regression | 6 | 86.4% | 0.30 | 0.97 | 0.87 | 8 | 26 | 6 |
| <sup>16</sup>CCM 2018 | Survival model | 4 | 67% | 0.85 | 0.67 | 0.85 | 16 | 30 | 19 |
| <sup>17</sup>JAMIA 2020 | RNN | 4 | — | 0.84 | 0.80 | 0.94 | 9 | 39 | 36 |
| <sup>18</sup>Nature Comm. 2021 | Random forest | 4 | — | 0.87 | 0.89 | 0.92 | 5 | 6 | 4 |
| <sup>10</sup>EMBC 2020 | GRU | 6 | 99.8% | 0.94 | 0.98 | 0.97 | 8 | 26 | 0 |
| <sup>19</sup>J. Elect.cardiology 2017 | Regression | 4 | 61% | 0.55 | 0.85 | 0.78 | 8 | 0 | 7 |
| <sup>20</sup>CIBM 2017 | LSTM | 3 | 93% | 0.94 | 0.91 | 0.93 | 8 | 0 | 1 |
| <sup>11</sup>ICHI 2018 | LSTM+CNN | 3 | 91.5% | 0.97 | 0.86 | 0.92 | 6 | 37 | 35 |
| **This work** | **Late fusion** | **4** | **90%**<sup>3</sup> | **0.90**<sup>3</sup> | **0.90**<sup>3</sup> | **0.90**<sup>3</sup> | **1** | **0** | **4** |
|  |  |  | **93%**<sup>4</sup> | **0.90**<sup>4</sup> | **0.95**<sup>4</sup> | **0.99**<sup>4</sup> | **1** | **0** | **5** |

<sup>1</sup>includes laboratory test results and culture results; <sup>2</sup>includes demographics and co-morbidities; <sup>3</sup>late fusion of demographics and ECG;
<sup>4</sup>late fusion of demographics, co-morbidity and ECG

### 3.2 Analog in-memory computation using switched-capacitor circuits

Energy efficiency of traditional AI computing systems are limited by communication costs of bringing together many input activations, and neuron weights, and distributing output activations in Von-Neumann architectures with separate memory and computing units. Wearable sensors have low energy budget which cannot accommodate conventional AI computing systems. IMC can break the Von-Neumann bottleneck by massively parallelizing computations and drastically reducing communication costs by performing computations using memory units. In this work, we perform analog IMC by using the capacitors that store the ANN weights to perform vector matrix multiplication. The complete vector matrix multiplications across all the neurons in each layer are performed simultaneously, and the results are stored locally in charge-domain on the shared top-plate of the capacitors. Vector matrix multiplication using capacitive IMC has high accuracy since arithmetic computation through passive charge sharing/redistribution is highly linear, and is less sensitive to random variations introduced during chip fabrication. Thanks to the analog IMC, the complete on-chip AI circuit consumes 7.4nJ/inference which is at least one order-of-magnitude lower than state-of-the-art medical ML ICs that consume hundreds of nJ to few µJ for inference^21–25^.

### 3.3 Limitations and future work

Our study has several limitations. First, the proposed architecture is only validated retrospectively on the dataset from a single institution - Emory University Hospital with high number of elderly patients (ICU records 2014-2018). Model trained on a single institution data may contain heavy bias towards the training population. We performed a race, gender, ethnicity and age based disparity analysis to understand the effect of bias using disparity measure of false positive rate (FPR) which is the fraction of false positives within labeled negatives of the group, and is critical for clinical event prediction (see Figure 4). Any disparity measure between 0.8 and 1.25 will be deemed fair which is inline with the 80 percent rule for determining disparity impact^?^. From the high-level overview, we found that many groups, specially for predictions 1 hour before sepsis onset, did not have equal or statistical parity, e.g. white population has low FPR disparity compared to black population, and patients aged over 61 have high FPR disparity compared to patients in the 41-60 age group. The disparity trend is similar but moderate for prediction 4 hours before sepsis onset. This disparity is primarily caused due to proportional disparity in the Emory ICU population. In our future work, we will design a clinical trial study to perform the validation on distinct patient populations. Second, requiring complete records for EMR and ECG would diminish the available patient pool to statistically irrelevant levels. However, within the bounds of limitation, we only used 6 most common data sources and our on-chip ECG model alone also achieved state-of-the-art performance for 4 hrs *t*_*onset*_. Third, while the capacitive analog IMC used in this work has the advantage of higher linearity and greater accuracy than other analog IMC techniques, such as using static random access memory (SRAM) arrays, a limitation of capacitive IMC in our work is that the AI model weights are hard-coded as capacitor values, and cannot be updated after chip fabrication. In our future work, we will use a novel analog IMC technique that combines high accuracy and linearity advantages of capacitive IMC with re-programmability of SRAM IMC by adding a capacitor to the SRAM cell which will allow vector matrix multiplication through passive charge sharing/redistribution while also allowing dynamic update of AI model weights. This will enable personalization of the AI models for each patient.

**Figure 4.**
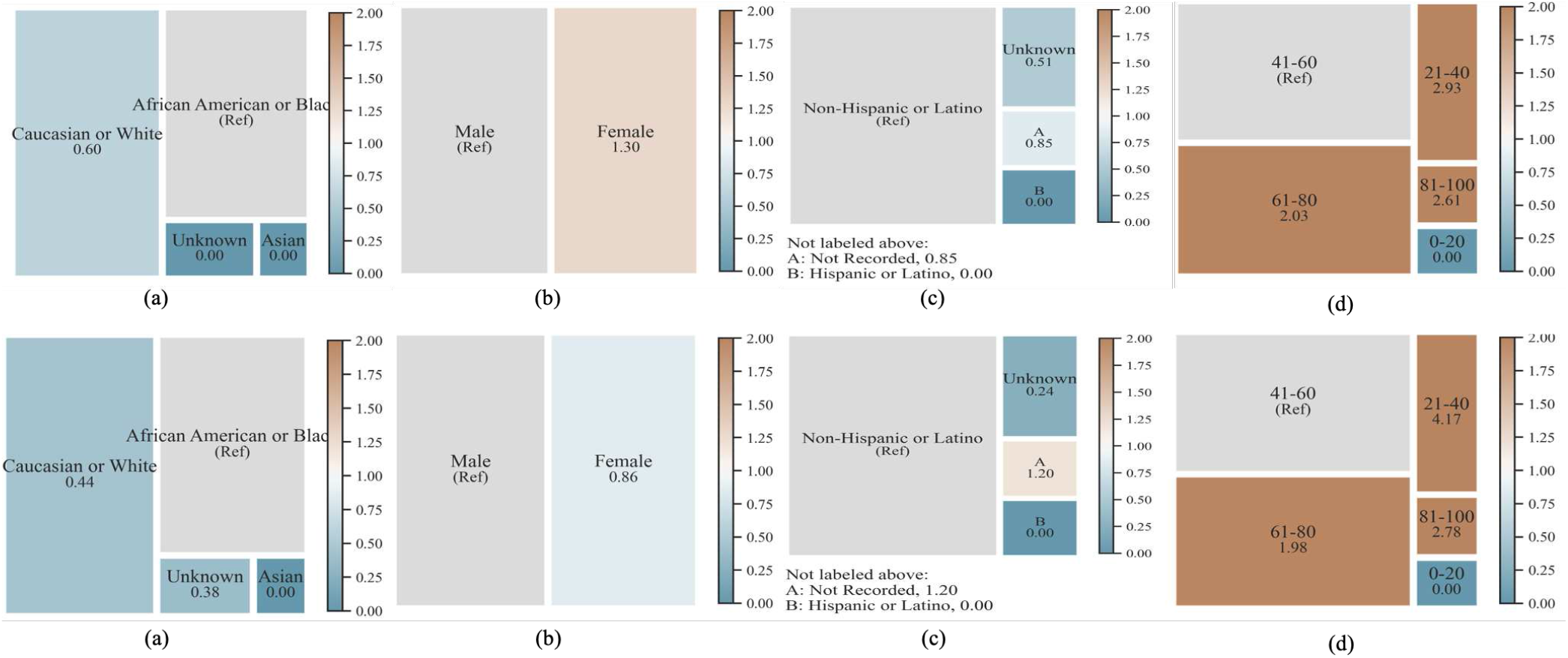
Disparity analysis results in terms of False Negative Rate (FNR) for late fusion using demographic, co-morbidity, and ECG data for different sepsis on-set prediction tasks; (First row) 1 hour; (Second row) 4 hours; (a) Race, (b) Gender, (c) Ethnic Group, (d)Age Category. Ref denotes reference value.

## 4 Methods

### 4.1 Dataset

With the approval of Emory Institutional Review Board (IRB), the de-identified sepsis dataset is obtained from Emory University Hospital (EUH). Following the Emory IRB regulation, we obtained waiver for informed consent since we used only de-identified data and no patient communication has been made during the study. The cohort consisted of 965 patients admitted to the ICUs at two hospitals within the Emory Healthcare system in Atlanta, Georgia from 2014 to 2018. For each patient, there is at least 8 hours of ECG signal recordings from the time of admission in the ICU, with the ECG signals sampled at 300Hz. Table 3 presents the overall patients demographics for both sepsis positive and negative patients. Our cohort mostly consist of patients older than 56 years. The Third International Consensus Definition of Sepsis (Sepsis-3) ^9^, criterion was used to assign sepsis onset time (tsepsis-3) when two conditions were simultaneously satisfied: 1) there was a clinical suspicion of infection (tsuspicion) and; 2) there was a two point increase in SOFA score (tSOFA). According to Sepsis-3 definition, the cohort consisted of a total of 965 patients, 514 of whom met the Sepsis-3 criterion.

**Table 3.** Patient characteristics table

| *Characteristics* |  | *Summary* |  |
| --- | --- | --- | --- |
|  |  | *Sepsis* | *Non-sepsis* |
| Data |  | 514 (53.26%) | 451 (46.73%) |
| Gender | Male | 281 (29.12%) | 214 (22.17%) |
|  | Female | 233 (24.14%) | 237 (24.56%) |
| Race | African American or Black | 191 (19.79%) | 190 (19.69%) |
|  | Caucasian or White | 278 (28.8%) | 215 (22.28%) |
|  | Asian | 8 (0.8%) | 7 (0.7%) |
|  | Hispanic | 1 (0.1%) | 0 (0%) |
|  | Multiple | 1 (0.1%) | 2 (0.2%) |
|  | American Indian or Alaskan Native | 0 (0%) | 2 (0.2%) |
|  | Unknown | 35 (3%) | 35 (3%) |
| Ethnic group | Hispanic or Latino | 9 (0.9%) | 7 (0.7%) |
|  | Non-Hispanic or Latino | 413 (42.78%) | 357 (36.99%) |
|  | Unknown | 92 (9.53%) | 87 (9.01%) |
| Age | 16-35 years | 52 (5.39%) | 42 (4.35%) |
|  | 36-55 years | 162 (16.79%) | 149 (15.44%) |
|  | 56 and above years | 300 (31.08%) | 260 (26.94%) |

### 4.2 On-chip ECG AI model

The ECG AI model is completely integrated within the analog sensor device. We designed a feature generation block that pre-processes the sensed signal by removing baseline wander, and then extract the pre-defined features. In the second stage, the extracted features are analyzed by the on-chip Artificial Neural Network (ANN) for the sepsis risk prediction.

#### 4.2.1 Pre-processing and feature extraction

ECG signal pre-processing and feature extraction are all performed on-chip, and hence, needs to be low-power without sacrificing accuracy. To remove baseline wander, we calculate median value of the ECG signal window and subtract it from all the samples in that window. For feature extraction, we consider only time-domain features based on first-order statistics of R-R peaks. While feature extraction in the literature is performed in both time-domain and frequency-domain, we choose only time-domain features since they are easy to compute on-chip at low power, compared to frequency-domain features which requires computation of frequency transform of the ECG signal. Frequency transform using fast fourier transform (FFT) is computationally expensive and consumes orders-of-magnitude higher power than time-domain features. We extract 14 time-domain features on 9000 samples ECG segments (30 seconds window). The extracted features are summarized in Figure 5 and are measures of central tendency, dispersion, shape of distribution of a window, R-R peaks, R-R intervals, variance between R peaks and average heart rate. The R peaks are identified in time-domain through thresholding. Figure 6(a) shows the training accuracy as a function of threshold used for identifying R peaks. The training accuracy drops if threshold is set too low or too high due to missing R peaks. The threshold for peak detection is set to 30% of the difference between maximum and minimum value for each segment.

**Figure 5.**
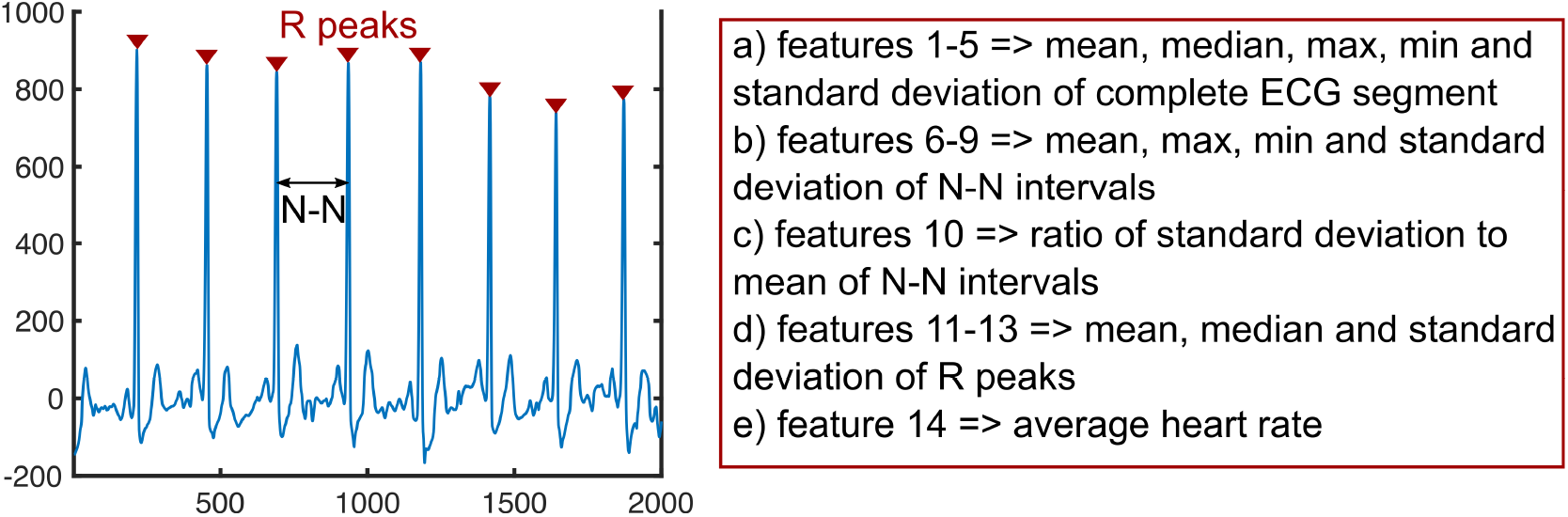
Summary of time-domain features extracted from ECG signal

**Figure 6.**
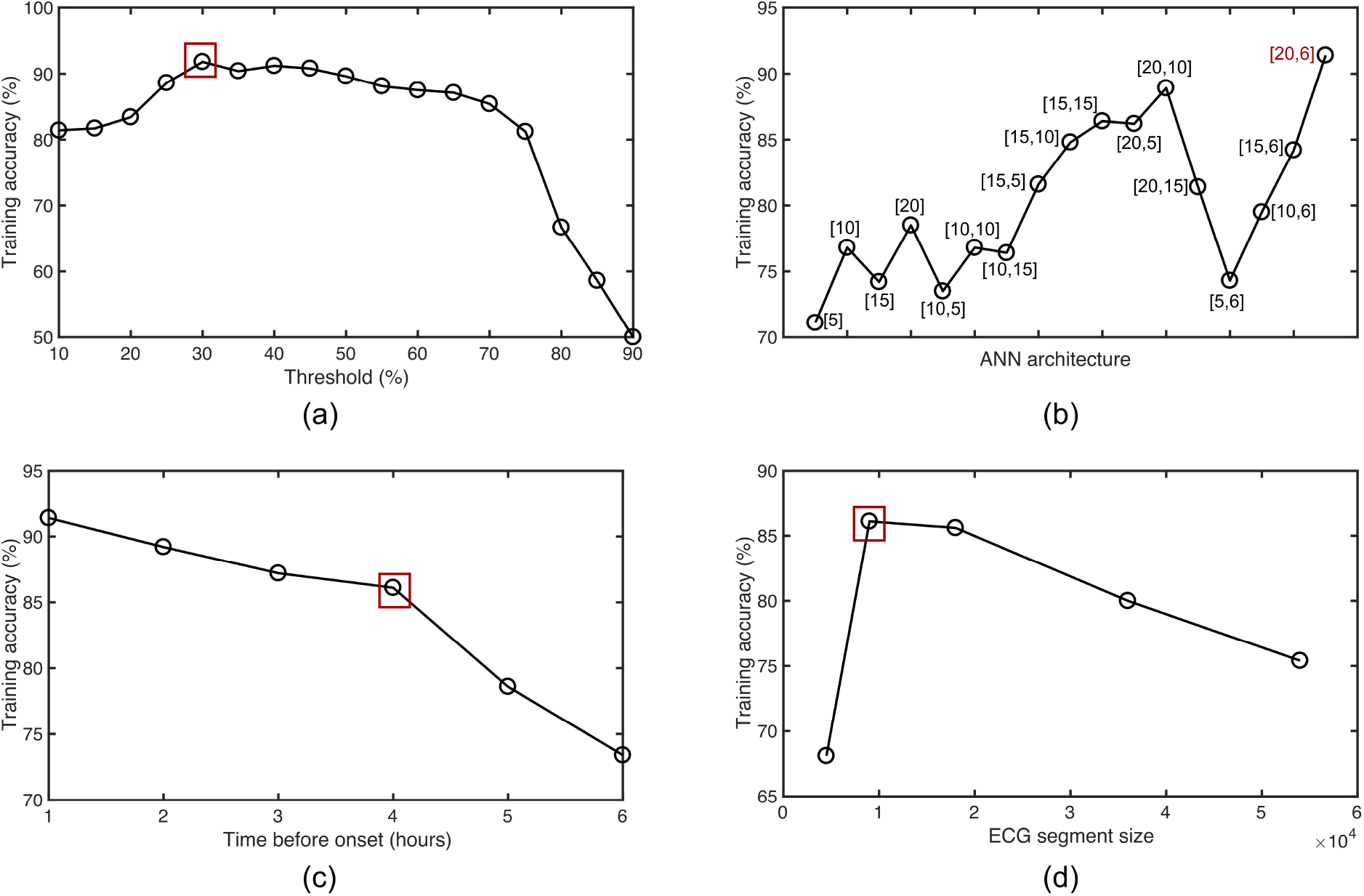
Training accuracy vs a) threshold for detecting R peaks b) ANN architecture c) time before sepsis onset d) ECG segment size

#### 4.2.2 Model training and hardware-software co-design

Figure 6(b) shows the training accuracy for different ANN architectures with the number of neurons in hidden layers annotated on the plot. The highest training accuracy is achieved for a 3-layer ANN with 20 neurons in the first hidden layer, and 6 neurons in the second hidden layer. The hidden layers use hyperbolic tangent (tanh) activation, while the output layer uses sigmoid activation. The dataset is partitioned randomly into 80% training data and 20% test data. Figure 6(c) shows the training accuracy as a function of time before sepsis onset. As is expected, the prediction accuracy drops monotonically with time before sepsis onset. In this plot, we present the prediction of sepsis 4 hours before onset. Figure 6(d) plots training accuracy versus size of ECG window in terms of sample size. The highest training accuracy is obtained for a segment size of 9000 samples corresponding to 30 seconds window.

Analog circuits are used to realize the ANN model in hardware. The digital features from the feature extractor are converted to analog form through 4-bit switched-capacitor digital-to-analog converters (DACs) before feeding to the analog ANN. The ANN performs multiply-and-accumulate (MAC) using switched-capacitors, and uses common-source differential amplifiers to realize tanh and sigmoid activations by leveraging intrinsic nonlinearity of transistors. Figure 7(a) and (b) shows the proposed custom tanh and sigmoid activations respectively, designed using 5 transistor, differential common-source amplifiers. Figure 7(c) shows the schematic of a neuron performing MAC using switched-capacitor circuits followed by analog activation. While a single-ended circuit is shown for simplicity, a fully differential architecture is used for each neuron. Compared to existing analog implementations which need 11-15 transistors for piece-wise approximation of ideal activation functions^26–29^, the proposed approach reduces transistor count, and hence, area. The proposed activation circuits use transistors biased in saturation region to minimize mismatch and noise. This is in contrast to existing analog techniques which bias transistors in sub-threshold to exploit exponential relation between drain current and gate voltage of transistors to implement tanh/sigmoid activation functions. Biasing transistors in sub-threshold region increases mismatch and reduces signal-to-noise ratio (SNR) compared to biasing in saturation region. Figure 7(d) shows layout of the complete ANN and feature extractor in 65nm CMOS process. The complete on-chip AI circuit consumes 7.4nJ/inference, out of which the feature extractor consumes 5.2nJ/inference, the 4-bit DACs consume 0.2nJ/inference, and the analog ANN consumes 2nJ/inference. Energy consumption of our on-chip AI circuit is at least one order-of-magnitude lower than state-of-the-art medical ML ICs that consume hundreds of nJ to few µJ for inference^21–25^.

**Figure 7.**
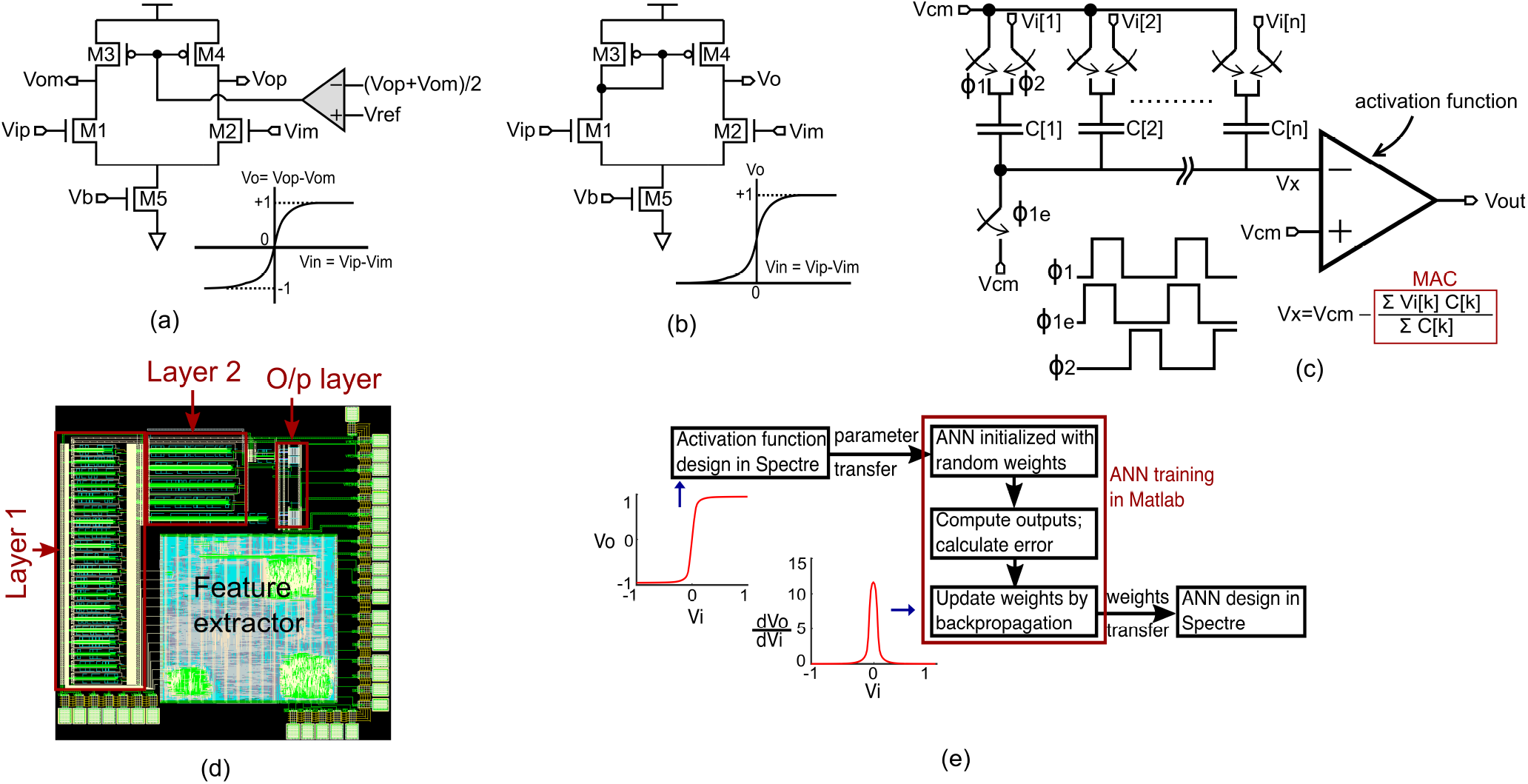
a) custom tanh activation circuit b) custom sigmoid activation circuit c) single-ended architecture of a neuron comprising switched-capacitor MAC and analog activation circuit d) layout of the complete sepsis classifier e) hardware-software co-design methodology

While the proposed activation functions reduce area and improve SNR, they approximate ideal mathematical functions, and, thus there are deviations between output of analog activation functions and mathematical counterparts. Instead of doing error correction at circuit level, which increases area and power cost, we use an error-aware ANN design methodology that takes into account the difference between ideal activation functions and the actual activation functions implemented using analog circuits to produce high accuracy AI predictions. In our prior study that focused on an image classification task^30,31^ we developed an error-aware hardware-software co-design interface in which SPICE simulations are used to characterize a unit activation function, and the characterization data is imported into Matlab for training the ANN (see Figure 7(e)). The co-simulation interface starts training with random weights for the ANN, creates SPICE netlist with the initial random weights and performs SPICE simulation of the ANN on the training dataset. The SPICE simulation outputs are parsed using Matlab and compared against validation and test output labels. Since the training is done with SPICE models of activation functions, rather than ideal mathematical functions, the discrepancies between software training and hardware implementation is minimized which allows use of analog circuits to approximate activation functions at low area and power cost.

### 4.3 EMR model

In addition to ECG signal data, we also incorporated the patient demographics and basic co-morbidities which are comparatively trivial to obtain at the patient side. In demographics, we considered four data elements - age, race, gender and ethnicity. Prior co-morbidities of the patients are coded as International Statistical Classification of Diseases and Related Health Problems (ICD) 10th revision. In order to maintain standardization and easy at-home use, we obtained mapping between disease names and ICD10 codes, and integrated the capability in the end-system to automatically map the disease name to the nearest ICD10 code. EMR data obtained underwent a series of pre-processing steps prior to formal analysis and model development.

Given the wide range of ICD10 codes (70,000 codes), we leverage the ICD10 disease description as string to obtained the embeddings of multiple comorbidities. The EMR data elements were both categorical (gender, age bins) and textual (ICD10 code descriptions) datatype. As the first data pre-processing step, we applied standard data cleaning steps, including removing empty cells, special characters. For the conversion of categorical features to numerical quantities, we use the label encoding technique. As per this technique, each value in a column is converted to a specific number.

The vectorization of ICD10 code descriptions was performed using Term Frequency-Inverse Document Frequency (Tf-idf) algorithm. Typically, the Tf-idf weight is composed by two terms: the first computes the normalized Term Frequency (TF), aka. TF(t) = (Number of times term t appears in a document) / (Total number of terms in the document); the second term is the Inverse Document Frequency (IDF), computed as the logarithm of the number of the documents in the corpus divided by the number of documents where the specific term appears-IDF(t) = log_*e*_(Total number of documents / Number of documents with term t in it). We trained the Tf-idf tokenizer using our training dataset and obtained 965 × 20 dimensional vector representation of the co-morbidities.

Finally the numeric representation of the categorical features and Tf-idf representation of the co-morbidities are combined using linear concatenation. We standardized features by removing the mean and scaling to unit variance. Centering and scaling happen independently on each feature by computing the relevant statistics on the samples in the training set. The mean (SD) is then used on later data (i.e, holdout test), using the same transformation function. The details of the patient characteristics have been shown in Table 3.

In order to perform a comprehensive analysis, we experimented with multiple parametric (logistic regression, Artificial Neural Network) and non-parametric (Linear Support Vector Machine - SVC, Random Forest) machine learning models. Given the static nature of this data, temporal sepsis onset prediction was not relevant and we only design a single prediction model for distinguishing sepsis vs non-sepsis data points using only EMR data. The optimal value of the hyperparameters is derived by analyzing the coefficient of the features computed by the 10-fold cross validation on the training data.

### 4.4 Fusion model

We hypothesize that an efficient combination of demographics along with co-morbidities, and multi-modal physiological signal data measured by our sensors will provide a more comprehensive view of a patient’s clinical status compared to an individual modality. We designed a prediction model that considers the feature vectors derived from continuous ECG data and integrate with patient demographics, commodities and baseline vitals. However the challenge was to integrate the static EMR features with the temporal physiological signal data in the same model. We designed a Late fusion model which performed a decision level fusion by leveraging predictions from multiple models (ECG model and EMR model) to make a final decision (Figure 8). In order to aggregate the prediction from multiple models, we trained a meta-learner to learn the importance of each model and design the aggregation function. For obtaining optimal performance, we experimented with four different classifier models as meta-learner - Linear SVC, Logistic Regression, Random Forest, and Neural Network with 2 hidden layers. We compared the late fusion model with the individual modalities in the Results section.

**Figure 8.**
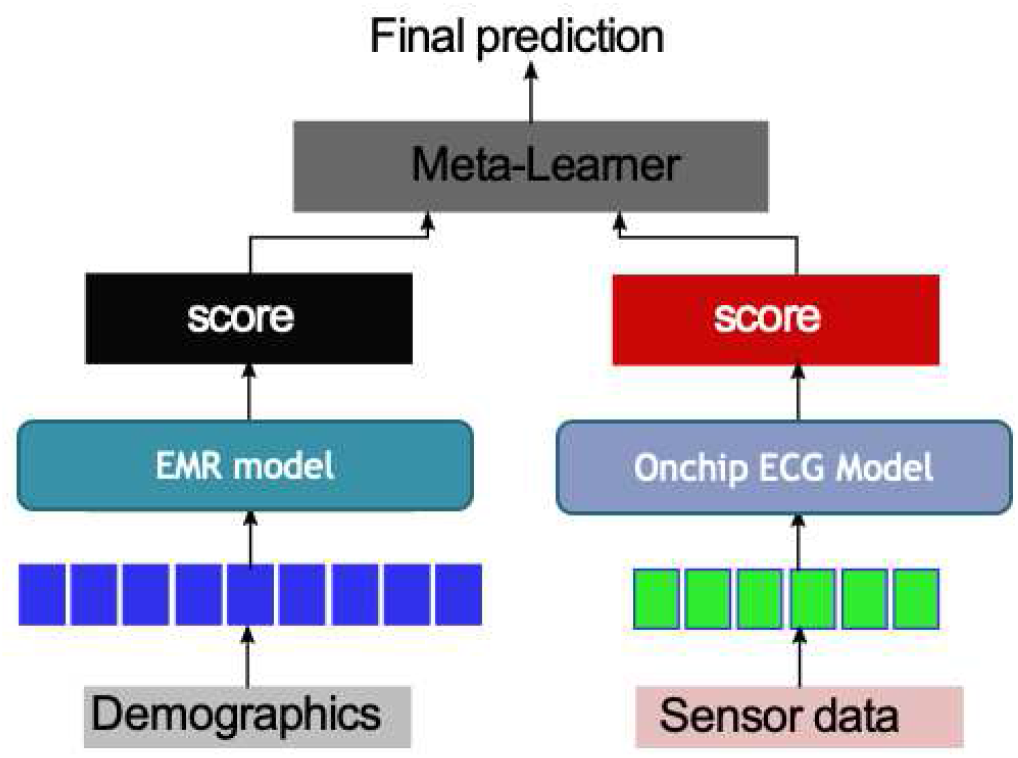
Late Fusion architecture with meta-learner - ECG and EMR model.

## Data Availability

Due to the HIPPA constrain the data cannot be shared with public.

## Acknowledgements

This material is based on research sponsored by Air Force Research Laboratory under agreement number FA8650-18-2-5402. The U.S. Government is authorized to reproduce and distribute reprints for Government purposes notwithstanding any copyright notation thereon. The views and conclusions contained herein are those of the authors and should not be interpreted as necessarily representing the official policies or endorsements, either expressed or implied, of Air Force Research Laboratory or the U.S. Government.

